# Safety and Immunogenicity of Intradermal Administration of Fractional Dose CoronaVac^®^, ChAdOx1 nCoV-19 and BNT162b2 as Primary Series Vaccination

**DOI:** 10.1101/2022.08.09.22278505

**Authors:** Somruedee Chatsiricharoenkul, Suvimol Niyomnaitham, H. Joshua Posen, Zheng Quan Toh, Paul V Licciardi, Patimaporn Wongprompitak, Thaneeya Duangchinda, Pattarakul Pakchotanon, Warangkana Chantima, Kulkanya Chokephaibulkit

## Abstract

There is a limited supply of COVID-19 vaccines, with less than 20% of eligible populations in low-income countries having received one dose. Intradermal delivery of fractional dose vaccines is one way to improve global vaccine access, but no studies have reported data on intradermal delivery of COVID-19 primary series vaccination. We conducted a pilot study to examine the safety and immunogenicity of three intradermal primary series regimens – heterologous regimen of CoronaVac and ChAdOx1 (CoronaVac-ChAdOx1), homologous regimen of ChAdOx1 (ChAdOx1-ChAdOx1), and homologous regimen of BNT162b2 (BNT162b2-BNT162b2). Each dose was 1/5^th^ or 1/6^th^ of the standard dose. Two additional exploratory arms of intradermal vaccination for the second dose following an intramuscular first dose of ChAdOx1 and BNT162b2 were included. Intradermal vaccination was found to be immunogenic and safe. The antibody responses generated by the intradermal primary series were highest in the BNT162b2 arms. The anti-receptor binding domain (anti-RBD) IgG concentration following fractional dose intradermal vaccination was similar to that of standard dose intramuscular vaccination of the same regimen, except for BNT162b2. The BNT162b2 intradermal series generated a lower antibody concentration than the reference intramuscular series, despite generating the highest antibody concentration of all three intradermal primary series regimens. Neutralizing antibody responses against the SARS-CoV-2 ancestral strain were consistent with what was observed for anti-RBD IgG, with lower titers for SARS-CoV-2 variants. The FRNT_50_ titers were lowest against the omicron variant, being undetectable (GMT≤10) in about a quarter of study participants. T-cell responses against spike- and nucleocapsid-membrane-open reading frame proteins were also detected following intradermal vaccination. Adverse effects following intradermal vaccination were generally comparable with post-intramuscular vaccination effects. Taken together, our data suggest that intradermal vaccination using 1/5^th^ or 1/6^th^ of standard COVID-19 intramuscular vaccination dosing generates similar immune responses with tendency of lower systemic adverse reactions than intramuscular vaccination. Our findings have implications in settings where COVID-19 vaccines are in shortage.

## Introduction

Intradermal vaccination has been touted as a possible solution to the insufficient supply of COVID-19 vaccines in resource-limited settings (1,2). Intradermal injection (ID) is known to stimulate a robust immune response for vaccination,(3) but is not commonly used because of the technical complexity of the procedure. The dermis has a higher concentration of antigen-presenting cells (APC) than exists in muscles or subcutaneous tissue (4). Consequently, a smaller dose of vaccine (10-20%) can induce similar immune responses when delivered ID compared with standard intramuscular (IM) or subcutaneous (SC) delivery (3). Further, ID delivery has been found to have fewer systemic adverse effects (1). The ID route is currently used for administration of Bacillus Calmette–Guérin and rabies vaccines.

Within the research landscape of intradermal delivery of COVID-19 vaccines, ChAdOx1 (manufactured by AstraZeneca) has been shown to stimulate dendritic cells and T cells (5,6), and a recent study in Thailand found a 20% fractional third (booster) dose of ChAdOx1 delivered ID, after a CoronaVac® (manufactured by Sinovac Life Sciences) 2-dose intramuscular primary series, was non-inferior to a standard IM booster dose of ChAdOx1 (7). Clinical trials are currently underway on ID delivery of primary series mRNA (BNT162b2 by Pfizer, and mRNA-1273 by Moderna) and viral vector vaccines (ChAdOx1) (8), and preliminary results have been encouraging. However, there are limited studies comparing ID and IM delivery of COVID-19 vaccines while the immunogenicity of ID delivery against emerging SARS-CoV-2 variants is unknown, making it difficult for policy makers to seriously consider ID vaccine delivery.

According to the national COVID-19 vaccination program of Thailand, the recommended primary series 2-dose regimens in 2021 were IM administration of heterologous CoronaVac-ChAdOx1, homologous ChAdOx1-ChAdOx1, and homologous BNT162b2-BNT-162b2. The immunogenicity of these regimens has been well studied (9,10). To address the gap in comparative data between IM and ID delivery routes, we conducted a pilot study to examine the safety and immunogenicity of ID delivery of the recommended primary series in Thailand, as well as the administration of a second dose via ID route following an IM first dose. We hypothesize that, across all permutations, ID delivery of fractional dose COVID-19 vaccines are immunogenic and has fewer systemic adverse effects. The findings of this study may guide policy decisions on vaccine distribution by decreasing the required vaccine supply per dose and increasing vaccine acceptability, thus resulting in increased vaccine coverage of the population and decreased COVID-19 related casualties.

## Materials and Methods

### Study design and subjects

This was a pilot single-center randomized prospective cohort study performed at a university-based referral center located in Bangkok, Thailand. This study aimed to examine the immunogenicity and safety of ID delivery of the three COVID-19 vaccine primary series regimens used in Thailand (CoronaVac-ChAdOx1, ChAdOx1-ChAdOx1 and BNT162b2-BNT162b2) and mixed delivery of IM-ID of homologous ChAdOx1 or BNT612b2 primary series. The study protocol was approved by the Siriraj Institutional Review Board (COA no. Si626/2021) and was registered with the Thai Clinical Trial Registry (TCTR20210903006).

Unvaccinated healthy participants aged 18 years or older were invited to join the study. Participants were excluded if they met any of the following criteria: a history of confirmed SARS-CoV-2 infection, positive SARS-CoV-2 serology, exposure within the prior 14 days to a COVID-19 patient without wearing adequate personal protective equipment, receipt of prophylactic treatment or investigational agents against COVID-19 within the prior 90 days, current immunocompromising medical condition, current use of an immunosuppressive agent, lifetime history of hypersensitivity to any vaccine, history of alcohol or drug abuse, regular cigarette smoking, underlying medical condition that may compromise the immune responses, or currently pregnant.

### Study procedures

Following written informed consent, participants were randomized to one of the five study arms of two-dose regimes: (1) ID CoronaVac-ID ChAdOx1 (N=20), (2) ID ChAdOx1-ID ChAdOx1 (N=20), (3) ID BNT162b2-ID BNT162b2 (N=20), (4):IM ChAdOx1-ID ChAdOx1 (N=10), and (5) IM BNT162b2-ID BNT162b2 (N=10). The mixed IM-ID mini-groups (arms 4 and 5) were designed to provide pilot data on the ID route when used as the second dose following primary ID (arms 2 and 3) or IM delivery. The volume of vaccine used for ID administration was 0.1mL of CoronaVac (1/5^th^ standard IM dose), 0.1 mL of ChAdOx1 (1/5^th^ standard IM dose), or 0.05mL of BNT162b2 (1/6^th^ standard IM dose). The interval between the two doses was 4±1 weeks in each study arm. Blood samples were collected at baseline, four weeks after first dose, and two and 12 weeks after the second dose for immunogenicity analysis.

Baseline blood samples were tested for anti-nucleoprotein antibody (anti-NP) using qualitative assay (Abbott, List No. 06R86) on the ARCHITECT I System, and anti-receptor binding domain (anti-RBD) IgG (see below) to exclude prior SARS-CoV-2 infection. Participants who were enrolled but later found to have positive anti-NP or anti-RBD at baseline were discontinued from the study.

Participants were observed for at least 30 minutes following vaccination to monitor for immediate adverse effects. They were then instructed to self-monitor for symptoms that could be considered adverse effects and submit an electronic diary (eDiary) entry using Google Forms® every day for 7 days after each vaccine dose. The eDiary solicited reporting of local adverse effects, including pain, erythema or swelling/induration at the injection site, localized axillary swelling, and tenderness anywhere along the arm that was injected. Systemic adverse effects solicited by the eDiary include headache, fatigue, myalgia, arthralgia, diarrhea, dizziness, nausea/vomiting, rash, fever, and chills. The severity of solicited adverse effects was graded using a numerical scale from 1 to 4 based on the Common Terminology Criteria for Adverse Events – Version 5.0 guide by the United States National Cancer Institute (NCI/NIH) (11).

### Humoral immune response

Humoral immune response was evaluated by measuring anti-RBD IgG to SARS-CoV-2 spike protein (ancestral strain) at all timepoints and neutralizing antibodies against ancestral and beta, delta and omicron variant strains at two weeks after second dose.

The anti-RBD IgG assay used the CMIA method of SARS-CoV-2 IgG II Quant (Abbott, List No. 06S60) on the ARCHITECT I System. This assay linearly measures the level of antibody between 21.0 – 40,000.0 arbitrary unit (AU)/mL, which was then converted to WHO International Standard concentration as binding antibody unit per mL (BAU/mL) following the equation provided by the manufacturer (BAU/mL = 0.142 x AU/mL). A level greater or equal to the cutoff value of 50 AU/mL or 7.1 BAU/mL was defined as seropositive.

Neutralizing antibodies against wuhan, delta, beta, and omicron variants were measured using the Focus Reduction Neutralization Test (FRNT) live virus method and calculated as the 50% inhibitory titer, or FRNT_50_. Procedures for FRNT were previously described (12). The Virus-infected cell foci were counted on the CTL-ImmunoSpot S6 Ultimate M2 analyzer using the ImmunoSpot software. The percentage of focus reduction was calculated using the probit program from the SPSS package. The titers of ≤10 were presented as 10.

### Cellular immune response

T-cell response was evaluated at four weeks after first dose and at two weeks after the second vaccine dose using human interferon gamma (IFN-γ) ELISpot kit (Mabtech, Nacka Strand, Sweden) according to manufacturer instructions. Briefly, peripheral blood mononuclear cells (PBMCs) were counted using an automated cell counter (Sysmex XN-10™Automated Hematology Analyzer). ELISpot plates were blocked for 30 min with 10% FCS containing RPMI media prior to the addition of 250,000 PBMCs/well. The stimulation solutions were S-peptide consisting of 100 peptides from spike protein, and NMO-peptide pools consisting of 101 peptides from nucleocapsid (N), membrane (M), open reading frame (ORF) 1, non-structural protein (nsp) 3, ORF-3a, ORF-7a, and ORF8 proteins. ELISpot plates were then incubated for 20 hours at 37°C and 5% CO2, washed and developed using a conjugated secondary antibody that bound to membrane-captured IFN-γ. The plates were read using IRIS (Mabtech) and spots were analyzed using Apex software 1.1 (Mabtech) and converted to spot-forming units (SFU) per million cells.

### Statistical Analysis

The anti-RBD IgG antibody responses were presented as geometric mean concentration (GMC) with 95% confidence intervals (CI). The FRNT_50_ data were presented as geometric mean titers (GMT) with 95% CIs. T-cell response as measured by geometric mean spot-forming units on ELISpot. The anti-RBD IgG GMCs of the same vaccine regimens delivered by standard IM route obtained from previously published study by our group using the similar laboratory facility were used as a reference (9,10) for the respective vaccine regimens delivered via ID in this study.

The seroconversion rates, anti-RBD IgG GMCs, and neutralizing antibody GMTs were compared within group and between the groups using paired t test and unpaired t test. The adverse effect endpoints were presented as frequencies (%) and compared using Fisher’s exact test. All statistical analyses were conducted using STATA version 17 (StataCorp, LP, College Station, TX, USA). P<0.05 is considered statistically significant.

## Results

### Study population

Between September and December 2021, 80 participants were recruited and randomized to one of the five study arms (Figure 1). Twenty participants were randomized to each of the study arms 1, 2 and 3, and 10 participants were randomized to each of study arms 4 and 5.

**Figure 1.**
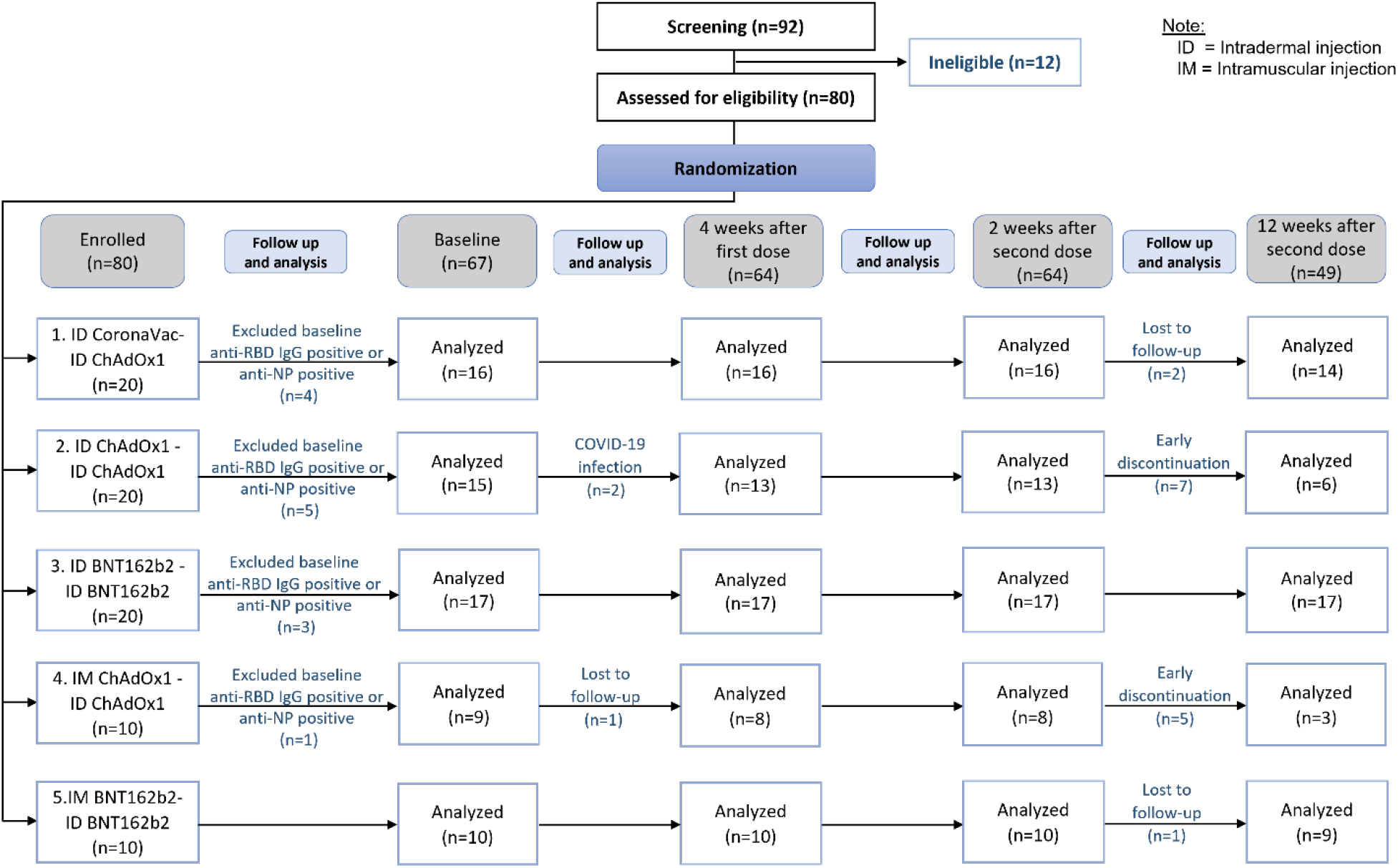
CONSORT subject flow diagram.

Thirteen participants were excluded from the analysis due to positive anti-RBD IgG or anti-NP IgG at baseline suggesting prior infection. Between the first and second dose, two participants from study arm two were infected with COVID-19 and excluded from further analyses. Seven and five participants in arm 2 and arm 4, respectively received additional vaccination between week 4 and 12 weeks follow up and therefore were excluded from the 12 weeks post-second dose analysis. Demographic characteristics including age, sex, and BMI were similar across all study arms (Table 1).

**Table 1.** Baseline characteristics of the subjects by routes and types of COVID-19 vaccines in first and second dose.

| First dose<br>-<br>Second dose | Routes and types of vaccines |  |  |  |  |  | P-value |
| --- | --- | --- | --- | --- | --- | --- | --- |
|  | All | ID<br>CoronaVac | ID<br>ChAdOx1 | ID<br>BNT162b2 | IM<br>ChAdOx1 | IM<br>BNT162b2 |  |
|  |  | ID<br>ChAdOx1 | ID<br>ChAdOx1 | ID<br>BNT162b2 | ID<br>ChAdOx1 | ID<br>BNT162b2 |  |
| Number of subjects (%) | 80<br>(100.0) | 20<br>(25.0) | 20<br>(25.0) | 20<br>(25.0) | 10<br>(12.5) | 10<br>(12.5) |  |
| Age (years); Median (IQR) | 34.5<br>(24.5,<br>44.0) | 36.5<br>(26.0,<br>45.5) | 35.0<br>(24.0,<br>41.5) | 32.5<br>(24.5,<br>42.5) | 36.5<br>(24.0,<br>48.0) | 29.5<br>(27.0,<br>36.0) | 0.928 |
| Male; n (%) | 41<br>(51.5) | 10<br>(50.0) | 12<br>(60.0) | 13<br>(65.0) | 3<br>(30.0) | 3<br>(30.0) | 0.218 |
| Body mass index (BMI: kg/m<sup>2</sup>);<br>Median (IQR) | 23.0<br>(20.4,<br>37.1) | 25.8<br>(20.3,<br>28.4) | 23.2<br>(20.9,<br>26.1) | 21.3<br>(18.6,<br>26.0) | 23.5<br>(22.5,<br>26.2) | 21.6<br>(20.6,<br>22.6) | 0.655 |
| Underweight<br>(< 18.50 kg/m<sup>2</sup>); n (%) | 9<br>(11.2) | 2<br>(10.0) | 1<br>(5.0) | 5<br>(25.0) | 1<br>(10.0) | 0<br>(0.0) | 0.117 |
| Normal weight<br>(18.50 - 24.90 kg/m<sup>2</sup>); n (%) | 44<br>(55.0) | 7<br>(35.0) | 14<br>(70.0) | 9<br>(45.0) | 6<br>(60.0) | 8<br>(80.0) |  |
| Overweight and obesity<br>(> 24.90 kg/m<sup>2</sup>); n (%) | 27<br>(33.7) | 11<br>(55.0) | 5<br>(25.0) | 6<br>(30.0) | 3<br>(30.0) | 2<br>(20.0) |  |

### Humoral immune response

A statistically significant increase in the GMC of anti-RBD IgG were observed across all study arms vaccine regimens, regardless of the vaccine or delivery method(s) four weeks after the first dose compared to baseline (Figure 2). Anti-RBD IgG peaked at two weeks after the second dose in all study arms, and thereafter declined by 12 weeks post second dose. Two weeks following second dose, regimens using BNT162b2 (study arms 3 and 5) had significantly higher peak GMC of anti-RBD IgG than the regimens using CoronaVac or ChAdOx1 (study arms 1, 2 and 4) (Figure 2). Compared with their respective IM-IM primary series, all study arms except study arm 3 (ID BNT162b2-ID BNT162b2) generated similar anti-RBD IgG at two weeks after the second dose, with trend towards lower GMC. Anti-RBD IgG at two weeks after the second dose in study arm 3 (ID BNT162b2-ID BNT162b2) were significantly lower (1.7-fold) than the IM BNT162b2-IM BNT162b2 reference (p=0.014), but the concentrations for study arm 3 were still significantly higher (at least 1.8-fold) than peak concentrations observed for study arm 1, 2 and 4 (p=0.017, p<0.001, and p<0.001, respectively).

**Figure 2.**
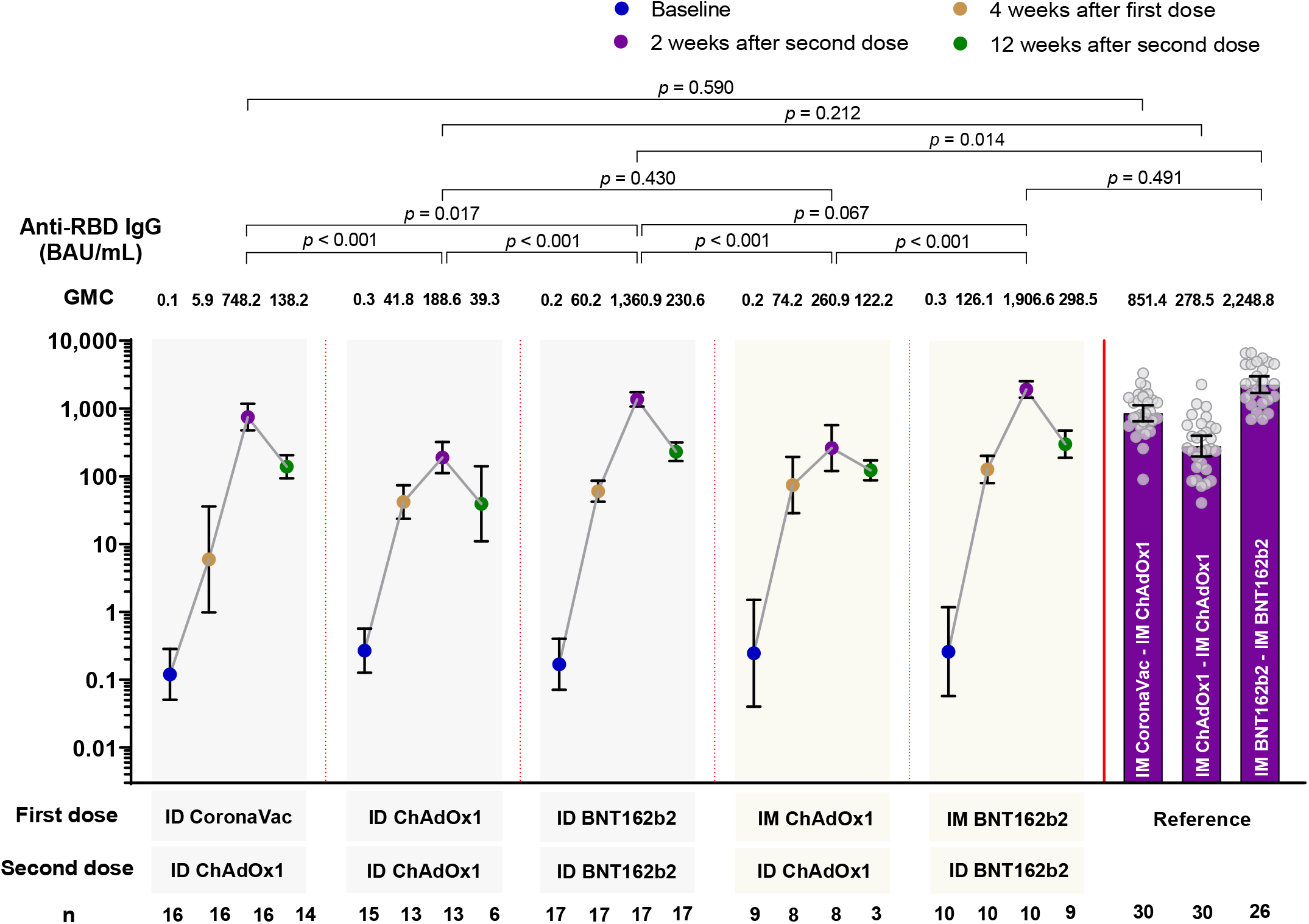
Geometric mean concentration of anti-RBD IgG at baseline, 4 months after first dose, 2 and 12 weeks after second dose for all study arms. Data from IM-IM vaccine regimens serve as the reference group for comparison (9). Data presented as geometric mean concentrations (GMC) and 95% confidence intervals. P<0.05 is considered statistically significant. BAU: binding antibody units. n= number of participants.

Comparing the anti-RBD IgG between the delivery methods of ID-ID and mix IM-ID (arm 2 versus arm 4, and arm 3 versus arm 5) at two weeks after second dose, lower concentrations were observed for ID-ID, but these were not statistically significant (p>0.05 for both comparisons) (Figure 2).

The GMT of neutralizing antibodies against four SARS-CoV-2 variants of each study arm are shown in Figure 3. Similar to anti-RBD IgG findings, study arms using BNT162b2 generated the highest GMT against all strains compared with regimens using ChAdOx1 and/or CoronaVac. The GMT of FRNT_50_ against the omicron variant was the lowest for all study arms compared with the ancestral strain, delta and beta variants. The proportion of subjects with titers against omicron ≤1:10 were 10/16 (62.5%), 13/13 (100%), 8/8 (100%), 12/17 (70.6%), and 4/10 (40%) arms 1, 2, 3, 4, and 5, respectively. Comparing the FRNT_50_ between the delivery methods of ID-ID and mix IM-ID (arm 2 versus arm 4, and arm 3 versus arm 5) at two weeks after second dose, lower FRNT_50_ were observed in ID-ID arms, but these were not statistically significant (p>0.05 for both comparisons).

**Figure 3.**
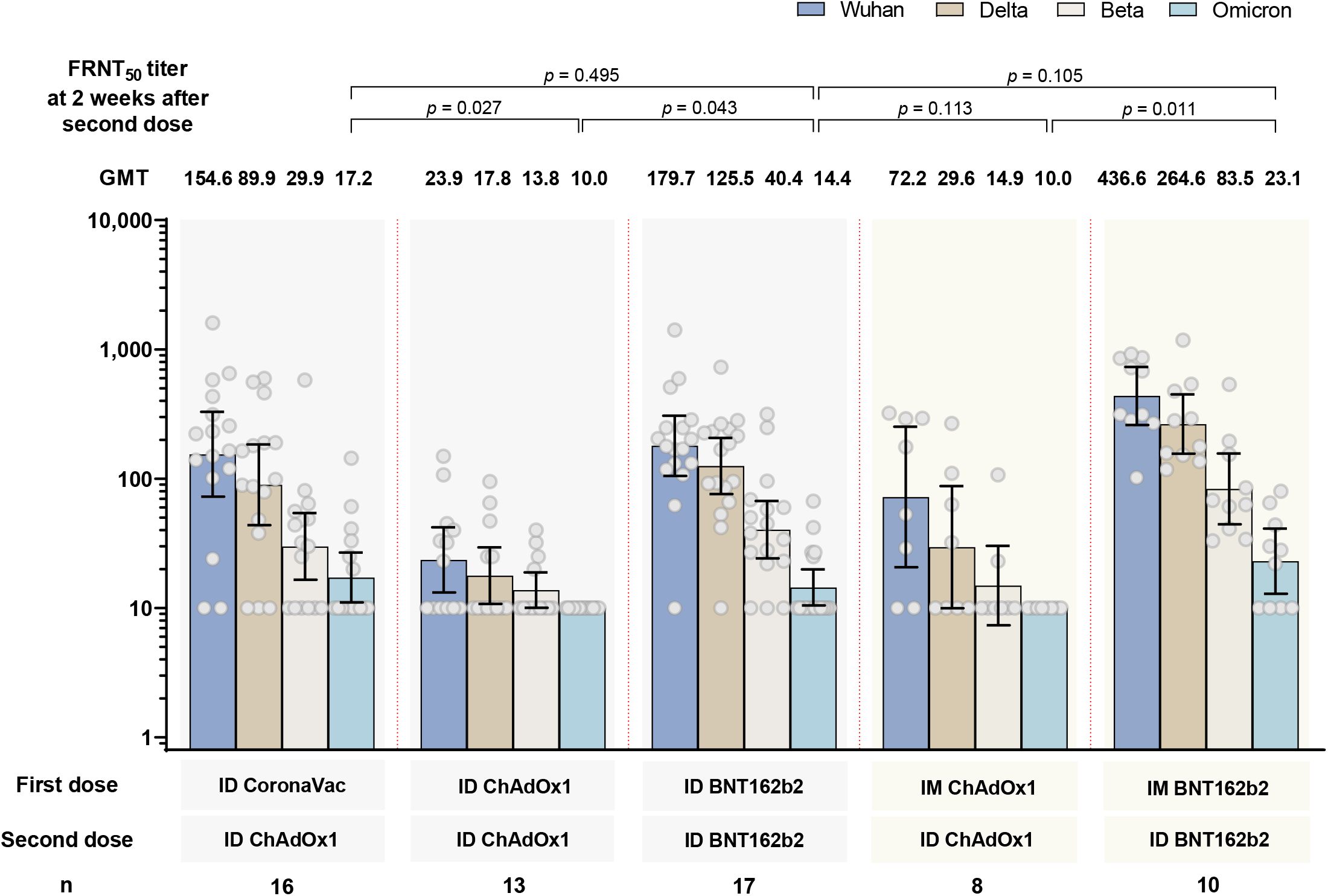
Neutralizing antibody titers (FRNT_50_) by focal reduction neutralization test against SARS-CoV-2 ancestral strain and beta, delta and omicron variant strains at 2 weeks after the second dose. Data presented as geometric mean concentrations and 95% confidence intervals. GMT: geometric mean titers. n= number of participants. ID: intradermal. IM: intramuscular.

### Cellular immune response

In general, there were no significant differences between ID-ID or IM-ID for ChAdOx1 and BNT162b2 study arms (Figure 4). ChAdOx1 given ID or IM induced the highest T-cell response against the ancestral Wuhan spike protein at four weeks after the first dose, and remained at similar level when measured at two weeks after the second dose. In contrast, while a lower spike-specific T cell response was observed at four weeks after the first dose of BNT162b2 (both IM and ID) compared to ChAdOx1, there was a significant increase in T cell response two weeks after second dose. ID CoronaVac-ID ChAdOx1 generated the lowest T cell response after two doses across the study arms, and the response was significantly lower than ID-ID BNT162b2 (Figure 4A). The T cell response against the ancestral NMO proteins were strongest in the ID CoronaVac group four weeks after the first dose (Figure 4B). However, there were no statistically significant differences between the study arms after dose 1 and dose 2, except for IM-ID ChAdOx1 which had significantly lower NMO T cell responses than IM-ID BNT162b2.

**Figure 4.**
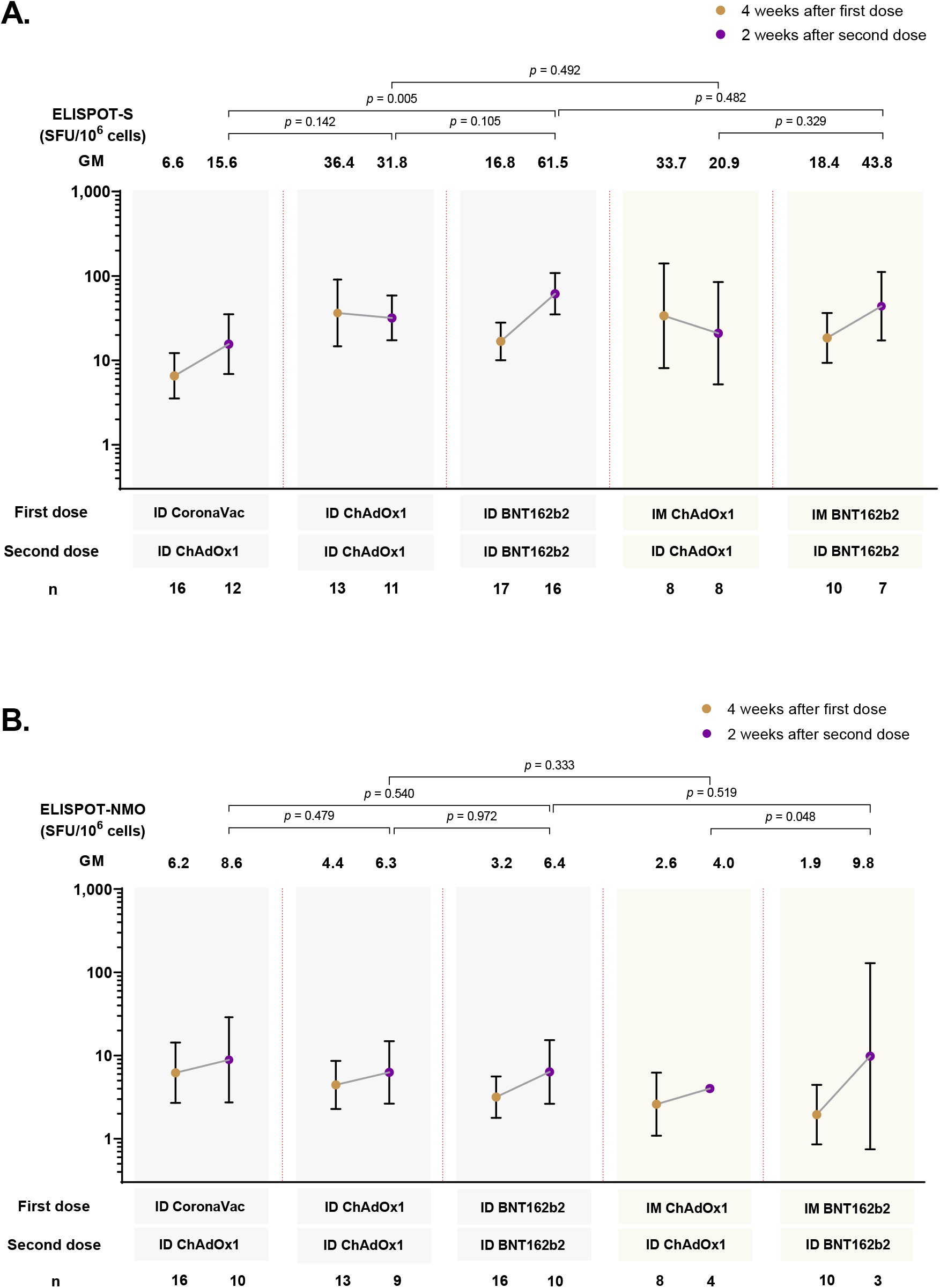
T-cell response against spike protein (A) and nucleocapsid-membrane-open reading frame (NMO) pool protein of ancestral Wuhan strain measured by ELISPOT at 4 weeks and 2 weeks after first and second vaccine dose, respectively. Data presented as geometric mean units (GMU) and 95% confidence intervals. n= number of participants. ID: intradermal. IM: intramuscular.

### Adverse reactions

Overall adverse reactions were common, but most were mild and only a small proportion of participants had moderate adverse reactions. On review of adverse reactions comparing the first dose of ID and IM of ChAdOx1 (arm 2 and 4), as well as of BNT162b2 (arm 3 and 5), there were fewer systemic events with ID delivery than with IM delivery but this was not significant (Figure 5A). In contrast, ID delivery tended to cause more local adverse reactions than IM delivery (Figure 5B). Arm 2 reported the highest frequency of local reaction with the average size of 2.31 cm. There were no major safety events.

**Figure 5.**
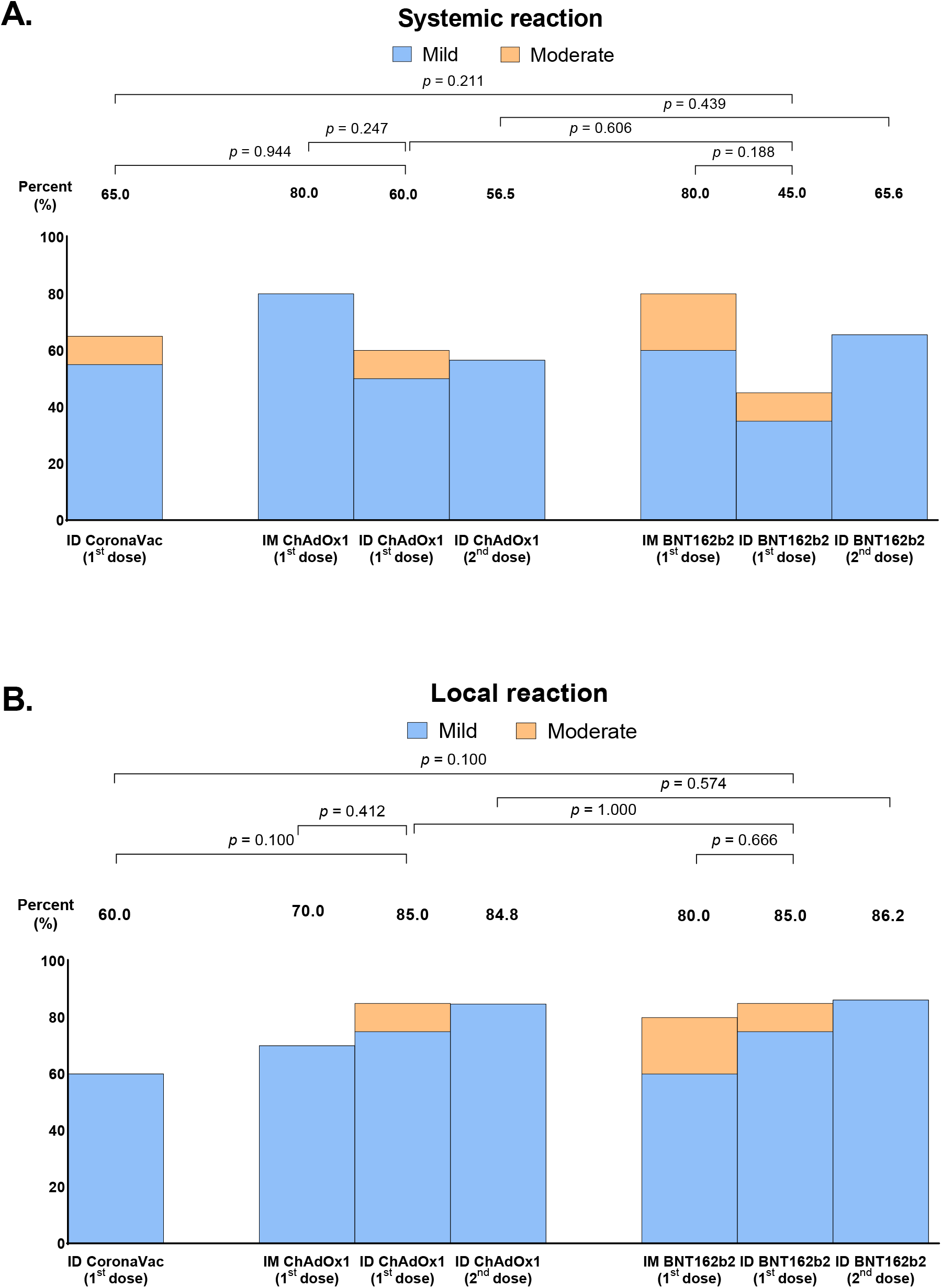
Systemic (A) and local adverse reactions (B) following intradermal or intramuscular-intradermal primary series. Data presented as percentage of individuals who reported any systemic or local adverse reactions.

## Discussion

To our knowledge, this is the first study to report on the immunogenicity and reactogenicity of intradermally delivered (ID-ID or IM-ID) 2-dose primary series of COVID-19 vaccination. Using only 1/5^th^ or 1/6^th^ dose of CoronaVac, ChAdOx1, or BNT162b2 delivered by ID injection, we found that regimens of heterologous CoronaVac-ChAdOx1, homologous ChAdOx1, and homologous BNT162b2, as well as a second dose given ID following an IM first dose for homologous ChAdOx1 or homologous BNT162b2 were immunogenic. These regimens using either one (IM-ID) or two ID (ID-ID) doses generated similar anti-RBD IgG responses as their respective standard IM-IM regimens, except for homologous BNT162b2 with both doses delivered ID, which induced a lower anti-RBD IgG concentration. Nonetheless, the anti-RBD IgG concentration generated by homologous BNT162b2 with two ID doses was still higher than that of ID or IM delivery of CoronaVac-ChAdOx1 or homologous ChAdOx1-ChAdOx1. Regarding reactogenicity, we found that ID delivery of these vaccines was well tolerated with mild but frequent local side effects, and trended towards lower systemic side effects compared to IM. Taken together, our data suggest that ID delivery of heterologous CoronaVac-ChAdOx1 or homologous ChAdOx1 or BNT162b2 as primary series or as second dose to IM delivery in the primary series may provide similar immunogenicity as IM delivery in the short term. The lower systemic reactogenicity may also improve vaccine uptake.

Our antibody data including the waning antibody responses up to three months following ID delivery is in line with previously reported data on the IM-IM primary series of CoronaVac, ChAdOx1 and BNT162b2, where BNT162b2 were most immunogenic compared with ChAdOx1 or CoronaVac (10). Also similar to prior studies on currently deployed IM regimens, we found that primary regimens using one or two doses of vaccine delivered via ID route had weak neutralizing antibodies against the omicron variant (13,14). The omicron variant has >60 mutations compared with the ancestral Wuhan strain, which increases its infectivity and ability to evade vaccine-induced immunity. A booster dose (3^rd^ vaccine dose) is needed to induce neutralizing antibodies and protect against the omicron variant (15). Administration of fractional doses of BNT162b2 as a booster via the ID route in individuals previously received CoronaVac or ChAdOx1 primary series via IM delivery of standard dose were found to induce lower neutralizing antibody responses against delta and omicron compared with the respective IM delivery (16), whereas ID delivery of ChAdOx1 to previously vaccinated CoronaVac individuals induced similar or higher neutralizing antibodies than corresponding IM delivery (17). Whether similar ID or IM boosting responses against SARS-CoV-2 is seen with individuals who received an ID primary series compared with individuals who received 3 doses of IM delivery, or 2 doses of IM and 1 booster dose of ID delivery remains to be determined. These results suggested the varied immunogenicity of different vaccine types when administering intradermally.

Our findings of T-cell response against the spike protein were similar to prior reports on IM vaccination with ChAdOx1 and BNT162b2 in a similar setting (18,19). This suggests that ID vaccination is may induce similar T cell responses as with IM. Interestingly, we found an increase in T-cell response against the NMO proteins in individuals vaccinated with BNT162b2. This observation needs to be interpreted with caution because of the small number of participants. This result may be true since increased antibodies to the N protein antigen have been documented in individuals vaccinated with spike-based mRNA vaccines (20). However, the clinical relevance of these findings is unknown, and whether a similar protection is offered by ID and IM vaccination remains to be determined.

Similar adverse reactions were found between ID and IM vaccination, but with a trend toward to lower systemic effects and slightly higher local effects following ID doses. This is consistent with what was reported for other vaccines (17,21,22). Lower systemic reaction from ID vaccination may be an advantage over IM vaccination, and may reduce vaccine hesitancy. A larger sample size would be needed to generate the statistical power needed to draw a clear conclusion.

The vaccine regimens evaluated in this study are currently implemented as IM regimens in Thailand and other low- or middle-income countries (LMIC). Our findings therefore have important implications for vaccine access in these countries. As of 22^nd^ July 2022, less than 20% of eligible population in low-income countries have received at least one dose of COVID-19 vaccine (23). Fractional dosage of ID vaccination using the current available vaccine as primary series may therefore help improve global vaccine coverage, provided similar protection can be achieved. Furthermore, our findings also provided a proof-of-concept for intradermal vaccination for new COVID-19 vaccines, including the omicron-specific mRNA vaccines that are currently under development or in clinical trials. These new vaccines are likely to be in shortage globally when they are first approved for use, and intradermal vaccination may alleviate this issue. However there is a need to consider for specific training on administration and supply of special syringes with intradermal vaccination.

Our study has some limitations. First, our sample size was small and insufficient to generate power to draw robust conclusions for the multiple comparisons. Second, we were not able directly compare our data with IM primary series in the same cohort and therefore our findings will need to be interpreted with caution. However, our IM comparison groups were based on published studies of the same setting and laboratory analysis in seronegative subjects, hence minimizing the potential variability (10). Third, we did not have a reference group for our neutralizing antibody and T cell analysis. Further studies are needed to confirm our findings. Finally, our data may not be generalizable to other COVID-19 vaccines.

In conclusion, we found that heterologous regimen of CoronaVac-ChAdOx1 and homologous ChAdOx1 or BNT162b2 delivered at a fractional (1/5^th^ or 1/6^th^) by ID was immunogenic. The results of ours and other previous studies on ID booster COVID-19 vaccination suggested that fractional dose of ID administration may induce similar antibody responses to standard IM administration for CoronaVac and ChAdOx1, but may be lower for BNT162b2. In addition, ID delivery has lower systemic adverse effects compared with standard dose IM delivery. Considering ID doses require only 10-20% the volume of IM doses, regimens incorporating ID doses should be considered as a possible partial solution to vaccine supply problems affecting LMICs and reduced concerns of vaccine safety and hesitancy. Our findings may also have implications for new COVID-19 vaccines in terms of decreasing the required vaccine supply per dose and increasing vaccine acceptability, thus resulting in increased vaccine coverage of the population and decreased COVID-19 related casualties.

## Data Availability

The raw data supporting the conclusions of this article are available by the corresponding authors upon request.

## Ethics statement

This study was approved by the Institutional Review Boards of the Committee on Ethics, Faculty of Medicine Siriraj Hospital, Mahidol University. (Certificate of approval Si626/2021)

## Author contributions

Conceptualization, SN, SC, KC; Methodology, SN, SC, PW, KC; Formal Analysis, SN, SC, HJP, ZQT, KC; Investigation, SN, SC, PW, TD, PP, WC, KC; Data Curation, SN; Writing – Original Draft Preparation, SC, ZQT, HJP, KC; Writing – Review & Editing, All authors.; Funding Acquisition, SC, SN, KC.

## Acknowledgments

We thank all the participants and the staff members who contributed to the study.

