## Supplementary figures and images for "Safety and Immunogenicity of Intradermal Administration of Fractional Dose CoronaVac^®^, ChAdOx1 nCoV-19 and BNT162b2 as Primary Series Vaccination"

### Supplementary Figure 1

Supplementary Figure 1

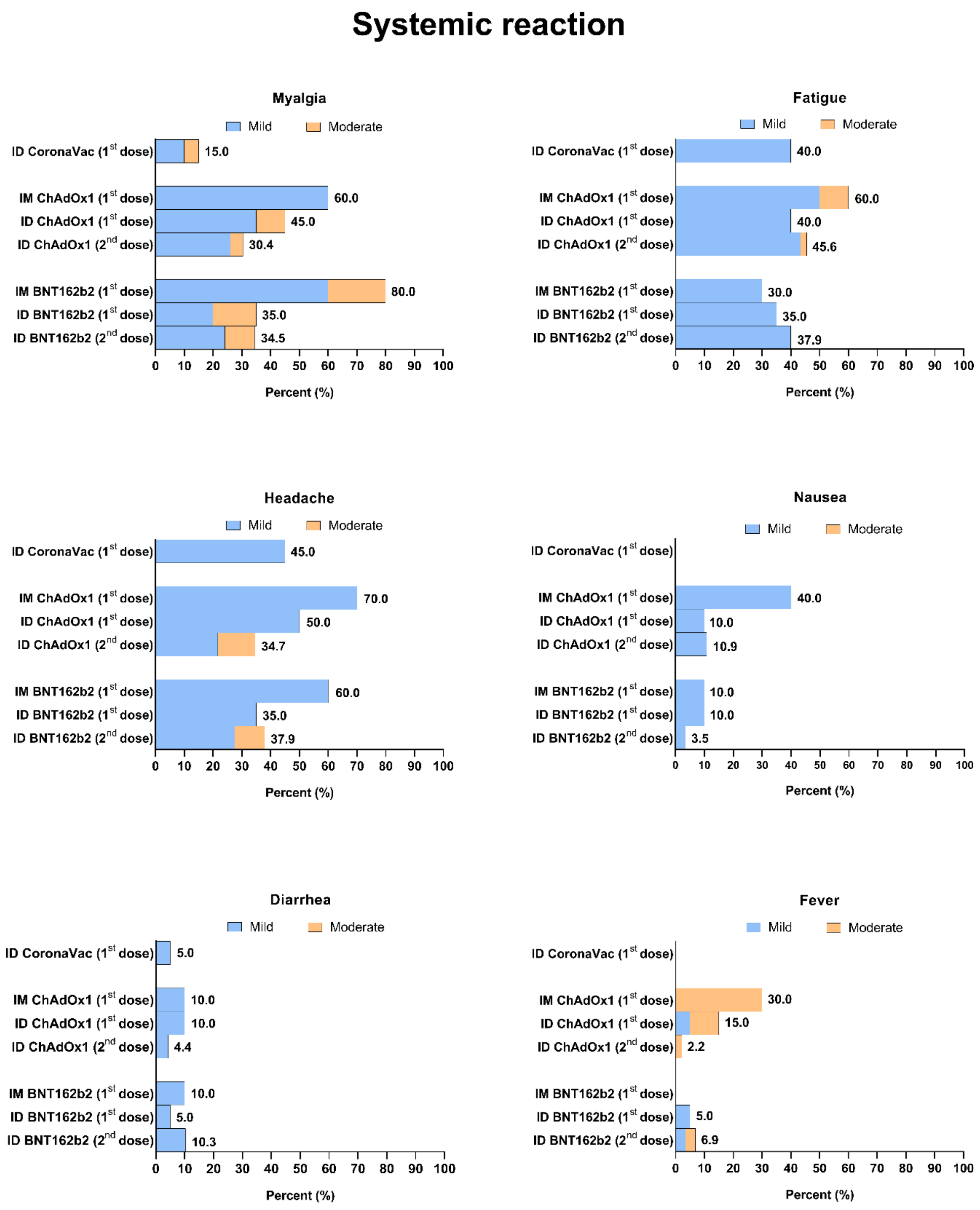
