## Supplementary Table 1 for "Safety and Immunogenicity of Intradermal Administration of Fractional Dose CoronaVac^®^, ChAdOx1 nCoV-19 and BNT162b2 as Primary Series Vaccination"

**Supplementary Table 1** Immunological response following routes and types of COVID-19 vaccines in first and second dose.

|  | **Routes and types of vaccines** | | | | | | |
| --- | --- | --- | --- | --- | --- | --- | --- |
| **First dose**  **-**  **Second dose** | **All** | **ID CoronaVac**  **- ID ChAdOx1** | **ID ChAdOx1**  **-**  **ID ChAdOx1** | **IM ChAdOx1**  **-**  **ID ChAdOx1** | **ID BNT162b2**  **-**  **ID BNT162b2** | **IM BNT162b2**  **-**  **ID BNT162b2** | ***p*-value** |
| **Enrolled** | **n=80** | **n=20** | **n=20** | **n=10** | **n=20** | **n=10** |  |
| Excluded | n=13 | Anti-np positive (n=3)  Anti-RBD positive  (n=1) | Anti-np positive  (n=3)  Anti-RBD positive  (n=2) | Anti-np positive (n=1) | Anti-np positive  (n=1)  Anti-RBD positive  (n=2) |  |  |
| **Anti-RBD IgG (BAU/mL)** | | | | | | | |
|  | **n=67** | **n=16** | **n=15** | **n=9** | **n=17** | **n=10** | ***p*-value** |
| GMC  at baseline (95% CI) | 0.2  (0.1, 0.3) | 0.1  (0.1, 0.3) | 0.3  (0.1, 0.6) | 0.2  (0.0, 1.5) | 0.8  (0.1, 0.4) | 0.3  (0.1, 1.2) | 0.701 |
|  | **n=64** | **n=16** | **n=13** | **n = 8** | **n = 17** | **n =10** | ***p*-value** |
| GMC  at 4 weeks after first dose  (95% CI) | 36.1  (21.4,  61.1) | 5.9  (0.9,  35.9) | 41.8  (23.7,  73.7) | 74.2  (28.5,  193.1) | 60.2  (42.2,  85.9) | 126.1  (79.2,  200.9) | <0.001 |
| GMR between 4 weeks after first dose / baseline  (95% CI) | 204.7  (111.6,  375.4) | 49.7  (8.7,  284.7) | 195.2  (85.6,  444.9) | 390.4  (67.3,  2265.4) | 356.1  (138.2,  917.9) | 487.3  (83.2,  2853.7) | 0.080 |
|  | **n=64** | **n=16** | **n=13** | **n=8** | **n=17** | **n=10** | ***p*-value** |
| GMC  at 2 weeks after second dose (95% CI) | 672.6  (508.7,  889.1) | 748.2  (475.7,  1176.9) | 188.6  (111.1,  320.2) | 260.9  (119.8,  568.0) | 1360.9  (1069.8,  17311.1) | 1906.6  (1441.6, 2521.7) | <0.001 |
| GMR between 2 weeks after second dose / baseline  (95% CI) | 3,811.9  (2323.0, 6255.3) | 6,249.9  (2283.3, 17107.1) | 881.4  (367.2,  2115.4) | 1,372.9  (260.2,  7244.8) | 8,050.7  (3389.2, 19123.4) | 7,366.1  (1572.5, 34505.5) | 0.005 |
| GMR between 2 weeks after second dose / 4 weeks after first dose (95% CI) | 18.6  (11.3,  30.7) | 125.8  (27.1,  584.1) | 4.5  (2.7,  7.5) | 3.5  (1.7,  7.4) | 22.6  (19.0,  26.9) | 15.1  (9.8,  23.3) | <0.001 |
|  | **n=49** | **n=14** | **n=6** | **n=3** | **n=17** | **n=9** | ***p*-value** |
| GMC  at 12 weeks after second dose (95% CI) | 161.8  (124.2,  210.7) | 138.2  (93.0,  205.4) | 39.3  (10.9,  140.6) | 122.2  (87.1,  171.3) | 230.6  (167.9,  316.7) | 298.5  (187.9,  474.0) | <0.001 |
| GMR between 12 weeks after second dose /baseline  (95% CI) | 994.3  (574.5, 1720.7) | 1,198.6  (363.8,  3948.5) | 244.7  (41.8,  1433.1) | 277.7  (2.7,  28960.5) | 1,364.2  (570.9,  3260.3) | 1593.1  (345.7,  7342.5) | 0.222 |
| GMR between 12 weeks / 2 weeks after second dose  (95% CI) | 0.2  (0.1, 0.2) | 0.2  (0.1, 0.2) | 0.2  (0.1, 0.3) | 0.2  (0.1, 0.8) | 0.2  (0.1, 0.2) | 0.2  (0.1, 0.2) | 0.888 |
| **Live virus focus reduction neutralization tests (FRNT_50_)** | | | | | | | |
|  | **n=64** | **n=16** | **n=13** | **n=8** | **n=17** | **n=10** | ***p*-value** |
| GMT against wuhan strain at 2 weeks after second dose  (95%CI) | 117.4  (81.4,  169.5) | 154.6  (72.7,  328.9) | 23.6  (13.2,  42.1) | 72.2  (20.7,  252.3) | 179.7  (105.3,  306.9) | 436.6  (260.8,  731.1) | <0.001 |
| GMT against delta strain at 2 weeks after second dose  (95%CI) | 72.8  (51.4,  103.1) | 89.9  (43.9,  184.1) | 17.8  (10.8,  29.5) | 29.6  (9.9,  87.9) | 125.5  (76.1,  206.9) | 264.6  (155.9,  449.1) | <0.001 |
| GMT against beta strain at 2 weeks after second dose  (95%CI) | 26.7  (18.5, 38.6) | 29.9  (16.5, 54.2) | 13.8  (10.1, 18.9) | 14.9  (7.4, 30.3) | 40.4  (24.2, 67.3) | 83.5  (44.5, 156.6) | <0.001 |
| GMT against omicron strain at 2 weeks after second dose  (95%CI) | 14.4  (12.2, 16.9) | 17.2  (11.1, 26.8) | 10.0  (10.0, 10.0) | 10.0  (10.0, 10.0) | 14.4  (10.5, 19.9) | 23.1  (12.9, 41.2) | 0.049 |
| Number (%) with FRNT_50_ against omicron strain ≤ 1:10 | 47  (73.4) | 10  (62.5) | 13  (100) | 8  (100) | 12  (70.6) | 4  (40.0) | 0.333 |
| GMR: wuhan/omicron strain at 2 weeks after second dose  (95%CI) | 8.2  (5.9, 11.1) | 8.9  (4.6, 17.7) | 2.4  (1.3, 4.2) | 7.2  (2.1, 25.2) | 12.5  (8.1, 19.1) | 18.9  (11.6, 30.9) | <0.001 |
| GMR: delta/omicron strain at 2 weeks after second dose  (95%CI) | 5.1  (3.8, 6.7) | 5.2  (2.9, 9.2) | 1.8  (1.1, 2.9) | 2.9  (0.9, 8.8) | 8.6  (5.7, 13.3) | 11.5  (8.1, 16.2) | <0.001 |
| GMR: beta/omicron strain at 2 weeks after second dose  (95%CI) | 1.9  (1.4, 2.5) | 1.7  (1.2, 2.5) | 1.4  (1.0, 1.9) | 1.5  (0.7, 3.0) | 1.9  (0.7, 5.4) | 3.6  (2.4, 5.4) | <0.001 |
| **ELISPOT responses (SU/mil)** | | | | | | | |
|  | **n=48** | **n=16** | **n=15** | - | **n=17** | - | ***p*-value** |
| ELISPOT-S  GMU at baseline (95% CI) | 3.0  (2.0, 4.5) | 1.9  (0.9, 3.6) | 3.1  (1.3, 7.4) | - | 4.6  (2.4, 8.9) | - | 0.172 |
| ELISPOT-NMO  GMU at baseline (95% CI) | 3.2  (2.2, 4.8) | 2.8  (1.4, 5.6) | 2.7  (1.1, 6.4) | - | 4.4  (2.3, 8.3) | - | 0.540 |
|  | **n=64** | **n=16** | **n=13** | **n=8** | **n=17** | **n=10** | ***p*-value** |
| ELISPOT-S  GMU at 4 weeks after first dose (95% CI) | 17.2  (12.2, 24.2) | 6.6  (3.5, 12.2) | 36.4  (14.6, 90.5) | 33.7  (8.1, 140.4) | 16.8  (10.0, 28.0) | 18.4  (9.3, 36.3) | <0.001 |
| ELISPOT-NMO  GMU at 4 weeks after first dose (95% CI) | 3.6  (2.6,  4.9) | 6.2  (2.7,  14.3) | 4.4  (2.3,  8.6) | 2.6  (1.1,  6.2) | 3.2  (1.8,  5.6) | 1.9  (0.9,  4.4) | 0.016 |
|  | **n=54** | **n=12** | **n=11** | **n=8** | **n=16** | **n=7** | ***p*-value** |
| ELISPOT-S  GMU at 2 weeks after second dose (95% CI) | 32.3  (22.9, 45.6) | 15.6  (6.9, 35.2) | 31.8  (17.3, 58.5) | 20.9  (5.2, 84.7) | 61.5  (35.0, 107.9) | 43.8  (17.2, 111.6) | 0.041 |
|  | **n=18** | **n=10** | **n=9** | **n=4** | **n=10** | **n=3** | ***p*-value** |
| ELISPOT-NMO  GMU at 2 weeks after second dose (95% CI) | 6.8  (4.9, 9.4) | 8.9  (2.7, 28.8) | 6.3  (2.6, 14.9) | 4.0  (4.0, 4.0) | 6.4  (2.6, 15.3) | 9.8  (0.8, 128.8) | 0.659 |

Note:	- p ≤ 0.05

- One-way ANOVA with parametric assumptions satisfied was determined *P*-value among those who received BNT162b2, ChAdOx1, and CoronaVac.

- Abbreviation: BAU/mL: binding antibody unit/mL, GMC: geometric mean concentration, GMT: geometric mean titer, GMU: geometric mean unit, GMR: geometric mean ratio, and SU/mil: spot forming unit per million cells
