## Supplementary Table 2 for "Safety and Immunogenicity of Intradermal Administration of Fractional Dose CoronaVac^®^, ChAdOx1 nCoV-19 and BNT162b2 as Primary Series Vaccination"

**Supplementary Table 2** Adverse events.

| **Adverse events** | **Routes and types of vaccines** | | | | | | |
| --- | --- | --- | --- | --- | --- | --- | --- |
| **First dose** | **All** | **ID CoronaVac** | **ID ChAdOx1** | **IM ChAdOx1** | **ID BNT162b2** | **IM BNT162b2** | ***p*-value** |
| **Number of subjects** | **n=80** | **n=20** | **n=20** | **n=10** | **n=20** | **n=10** |  |
| **After first dose** | | | | | | | |
| **Local reaction, n (%)**  Mild, n (%)  Moderate, n (%) | 61 (76.3)  55 (68.7)  6 (7.5) | 12 (60.0)  12 (60.0)  0 (0.0) | 17 (85.0)  15 (75.0)  2 (10.0) | 7 (70.0)  7 (70.0)  0 (0.0) | 17 (85.0)  15 (75.0)  2 (10.0) | 8 (80.0)  6 (60.0)  2 (20.0) | <0.001 |
| **Systemic reaction, n (%)**  Mild, n (%)  Moderate, n (%) | 50 (62.5)  42 (52.5)  8 (10.0) | 13 (65.0)  11 (55.0)  2 (10.0) | 12 (60.0)  10 (50.0)  2 (10.0) | 8 (80.0)  8 (80.0)  0 (0.0) | 9 (45.0)  7 (35.0)  2 (10.0) | 8 (80.0)  6 (60.0)  2 (20.0) | <0.001 |
| **Myalgia, n (%)**  Mild, n (%)  Moderate, n (%) | 33 (41.3)  24 (30.0)  9 (11.3) | 3 (15.0)  2 (10.0)  1 (5.0) | 9 (45.0)  7 (35.0)  2 (10.0) | 6 (60.0)  5 (50.0)  1 (10.0) | 7 (35.0)  4 (20.0)  3 (15.0) | 8 (80.0)  6 (60.0)  2 (20.0) | 0.103 |
| **Fatigue, n (%)**  Mild, n (%)  Moderate, n (%) | 32 (40.0)  31 (38.7)  1 (1.3) | 8 (40.0)  8 (40.0)  0 (0.0) | 8 (40.0)  8 (40.0)  0 (0.0) | 6 (60.0)  5 (50.0)  1 (10.0) | 7 (35.0)  7 (35.0)  0 (0.0) | 3 (30.0)  3 (30.0)  0 (0.0) | 0.619 |
| **Headache, n (%)**  Mild, n (%)  Moderate, n (%) | 39 (48.8)  39 (48.8)  0 (0.0) | 9 (45.0)  9 (45.0)  0 (0.0) | 10 (50.0)  10 (50.0)  0 (0.0) | 7 (70.0)  7 (70.0)  0 (0.0) | 7 (35.0)  7 (35.0)  0 (0.0) | 6 (60.0)  6 (60.0)  0 (0.0) | 0.763 |
| **Nausea, n (%)**  Mild, n (%)  Moderate, n (%) | 9 (11.3)  9 (11.3)  0 (0.0) | 0 (0.0)  0 (0.0)  0 (0.0) | 2 (10.0)  2 (10.0)  0 (0.0) | 4 (40.0)  4 (40.0)  0 (0.0) | 2 (10.0)  2 (10.0)  0 (0.0) | 1 (10.0)  1 (10.0)  0 (0.0) | 0.091 |
| **Diarrhea, n (%)**  Mild, n (%)  Moderate, n (%) | 6 (7.5)  6 (7.5)  0 (0.0) | 1 (5.0)  1 (5.0)  0 (0.0) | 2 (10.0)  2 (10.0)  0 (0.0) | 1 (10.0)  1 (10.0)  0 (0.0) | 1 (5.0)  1 (5.0)  0 (0.0) | 1 (10.0)  1 (10.0)  0 (0.0) | 0.983 |
| **Fever, n (%)**  Mild, n (%)  Moderate, n (%) | 7 (8.7)  2 (2.5)  5 (6.2) | 0 (0.0)  0 (0.0)  0 (0.0) | 3 (15.0)  1 (5.0)  2 (10.0) | 3 (30.0)  0 (0.0)  3 (30.0) | 1 (5.0)  1 (5.0)  0 (0.0) | 0 (0.0)  0 (0.0)  0 (0.0) | 0.134 |
| **Second dose** | **Total** | **ID ChAdOx1** | **ID BNT162b2** |  | | | ***p*-value** |
| **Number of subjects** | **n=75** | **n=46** | **n=29** |  |  |  |  |
| **After second dose** | | | |  |  |  |  |
| **Local reaction, n (%)**  Mild, n (%)  Moderate, n (%) | 64 (85.3)  64 (85.3)  0 (0.0) | 39 (84.8)  39 (84.8)  0 (0.0) | 25 (86.2)  25 (86.2)  0 (0.0) |  | | | 0.055 |
| **Systemic reaction, n (%)**  Mild, n (%)  Moderate, n (%) | 45 (60.0)  45 (60.0)  0 (0.0) | 26 (56.5)  26 (56.5)  0 (0.0) | 19 (65.6)  19 (65.6)  0 (0.0) |  |  |  | <0.001 |
| **Myalgia, n (%)**  Mild, n (%)  Moderate, n (%) | 24 (32.0)  19 (25.3)  5 (6.7) | 14 (30.4)  12 (26.0)  2 (4.4) | 10 (34.5)  7 (24.1)  3 (10.3) |  |  |  | <0.001 |
| **Fatigue, n (%)**  Mild, n (%)  Moderate, n (%) | 32 (42.6)  31 (41.3)  1 (1.3) | 21 (45.6)  20 (43.4)  1 (2.2) | 11 (37.9)  11 (37.9)  0 (0.0) |  |  |  | <0.001 |
| **Headache, n (%)**  Mild, n (%)  Moderate, n (%) | 27 (36.0)  18 (24.0)  9 (12.0) | 16 (34.7)  10 (21.7)  6 (13.0) | 11 (37.9)  8 (27.5)  3 (10.4) |  |  |  | <0.001 |
| **Nausea, n (%)**  Mild, n (%)  Moderate, n (%) | 6 (8.0)  6 (8.0)  0 (0.0) | 5 (10.9)  5 (10.9)  0 (0.0) | 1 (3.5)  1 (3.5)  0 (0.0) |  |  |  | 0.716 |
| **Diarrhea, n (%)**  Mild, n (%)  Moderate, n (%) | 5 (6.7)  5 (6.7)  0 (0.0) | 2 (4.4)  2 (4.4)  0 (0.0) | 3 (10.3)  3 (10.3)  0 (0.0) |  |  |  | 0.926 |
| **Fever, n (%)**  Mild, n (%)  Moderate, n (%) | 3 (4.0)  1 (1.3)  2 (2.7) | 1 (2.2)  0 (0.0)  1 (2.2) | 2 (6.9)  1 (3.5)  1 (3.4) |  |  |  | <0.001 |

Note:	Abbreviation: ID = Intradermal injection, IM = Intramuscular injection, and IQR = Interquartile range
